# From more testing to smart testing: data-guided SARS-CoV-2 testing choices

**DOI:** 10.1101/2020.10.13.20211524

**Authors:** Janko van Beek, Zsofia Igloi, Timo Boelsums, Ewout Fanoy, Hannelore Gotz, Richard Molenkamp, Jeroen van Kampen, Corine GeurtsvanKessel, Annemiek van der Eijk, David van de Vijver, Marion Koopmans

## Abstract

We present an in-depth analysis of data from drive through testing stations using rapid antigen detection tests (RDT’s), RT-PCR and virus culture, to assess the ability of RDT’s to detect infectious cases. We show that the detection limits of five commercially available RDT’s differ considerably, impacting the translation into the detection of infectious cases. We recommend careful fit-for-purpose testing before implementation of antigen RDT’s in routine testing algorithms as part of the COVID-19 response.

## Main text

Rapid detection and isolation of new cases, combined with contact tracing and quarantine has become a critical pillar of the global efforts to reduce circulation of SARS-CoV-2. The purpose of this test, trace and isolate (TTI) strategy is to stop transmission chains and reduce the impact of COVID-19. Epidemiological modelling suggests that aggressive TTI combined with physical distancing and the use of personal protective equipment when physical distancing is not achievable, could suppress a second wave without the need for a prolonged lockdown^1^.

The case numbers have increased in recent weeks across Europe (https://covid19-country-overviews.ecdc.europa.eu/) and the capacity to keep up the TTI strategy is reaching its limits. Therefore, there is an urgent need for optimising the available resources. Currently, case diagnosis relies on RT-PCR testing, which has limitations in terms of time to result, and for which scaling up capacity has been hampered by scarcity of critical reagents. In addition, while highly specific, the sensitivity of RT-PCR combined with prolonged shedding of low amounts of viral RNA for weeks may lead to positive test results following clinical recovery long after a person is infectious^2^. Furthermore, screening of persons without symptoms may yield weak positive test results, raising questions about how to handle such cases. Ideally, screening of cases would be based on testing for infectivity, but such cell culture based assays have long turnaround times and therefore are not suitable for rapid screening, which is essential for the success of TTI. Rapid antigen detection tests (RDT) have recently entered the diagnostic market. Compared with RT-PCR, they are relatively easy to produce, cheaper, easy to use with faster turn-around times, and, depending on the assay, without the need for dedicated equipment or high level laboratory capacity. The widespread and frequent use of such tests has recently been proposed as a solution to the safe reopening and return to the pre-pandemic social interactions, but to our knowledge there is no data about the performance in detecting infectious cases^3^.

Here, we assess the potential impact of introduction of RDT’s in the current test strategy of the Netherlands in which the majority of testing is done in drive through test stations. Detection limits of 5 commercially available RDT’s were determined using serial dilutions of freshly harvested SARS-CoV-2 virus stock. Freshly collected nasal and nasopharyngeal samples in viral transport media from people presenting to the drive through test station with a range of Ct values were tested in parallel by RT-PCR, and RDT. Samples were also inoculated onto Vero E6 cells to assess the correlation between RT-PCR/antigen rapid test results and infectiousness for persons with mild symptoms, to supplement published data on this relationship for hospitalised patients and patients with mild symptoms^2,4^.

Quantitative data on the distribution of viral RNA loads for all cases tested positive at the test location since June 1st (N = 1754), results of RT-PCR testing, expressed as cycle threshold (Ct) values were retrieved from the laboratory database for the two different RT-PCR platforms used. To allow direct comparison of results, the Ct values were translated into copy numbers by determining the amount of RNA copies based on the E-gene RT-PCR and quantified E-gene *in vitro* RNA transcripts as described by Corman *et al*. and using an in-house prepared standard of cultured virus^5^. For both platforms, turnaround time was logged systematically and this data was extracted to assess potential impact of implementation of antigen testing on the time to result. To determine the profile of positive patients in the drive-through test stations, we selected data from 223 COVID-19 index cases with complete background on date of symptom onset, date of sample collection date of diagnosis from the database of the municipal health service of Rotterdam-Rijnmond.

The distribution of Ct values and estimated RNA loads for patients reporting to drive through testing was skewed to high RNA loads (Figure 1). Data on time since symptom onset for a selection of cases showed that the majority of persons report to the testing station within the first week of symptom onset, in line with national recommendations for testing (Figure 2). The viral load distribution for the complete patient group (N = 1754) was similar, and therefore we assumed a similar profile of cases (data not shown). Results were available the same day for 33% of cases, the next day for 55%, after 2 days for 11% and after 3 days for 1%. Infectiousness was assessed by including the matched viral cultures and RT-PCR of 78 randomly selected individuals that were diagnosed with SARS-CoV2 in the drive through testing station. We determined the probability of being infectious for all patients tested in the drive through station using logistic regression analysis in R 4.0.0. This analysis was used to calculate the density distribution of infectious persons (Figure 2, red bars). In addition, infectiousness was calculated based on two published studies that tested cell culture in parallel with RT-PCR in hospitalized patients with severe and mild illness, respectively (Van Kampen *et al*., 2020; Wölfel *et al*., 2020) (Supplementary figure 1). This comparison suggests that throat/nose swabs from outpatients more often were cell culture positive than those from hospitalised cases.

**Figure 1.**
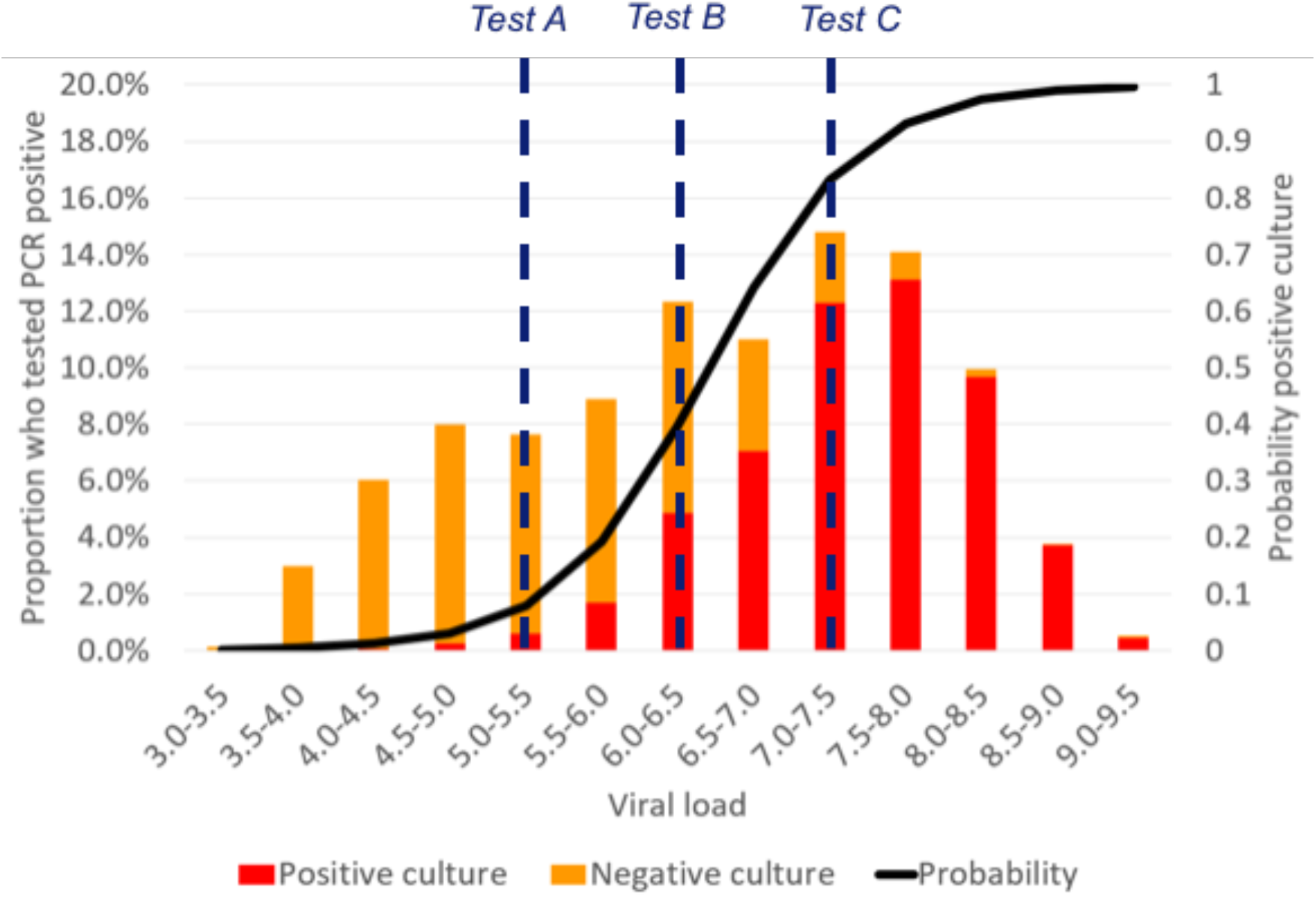
Distribution of viral RNA loads at time of diagnosis for n=1754 patients with RT-PCR confirmed SARS-CoV-2 infection presenting to a drive through test station (orange bars), and distribution of patients shedding infectious virus at time of diagnosis (red bars). Black line denotes a logistic regression curve of probability of having infectious virus in the nose/throat swab. Dashed vertical lines indicate the technical detection limits of the different rapid tests. Assay A = *Panbio™ COVID-19 Ag rapid test (Abbott)*, and Standard Q COVID-19 Ag (SD Biosensor); Assay B = *COVID-19 Ag Respi-Strip (Coris BioConcept)*, and GenBody COVID-19 Ag (GenBody Inc); Assay C = Biocredit COVID-19 Ag (RapiGEN).

**Figure 2.**
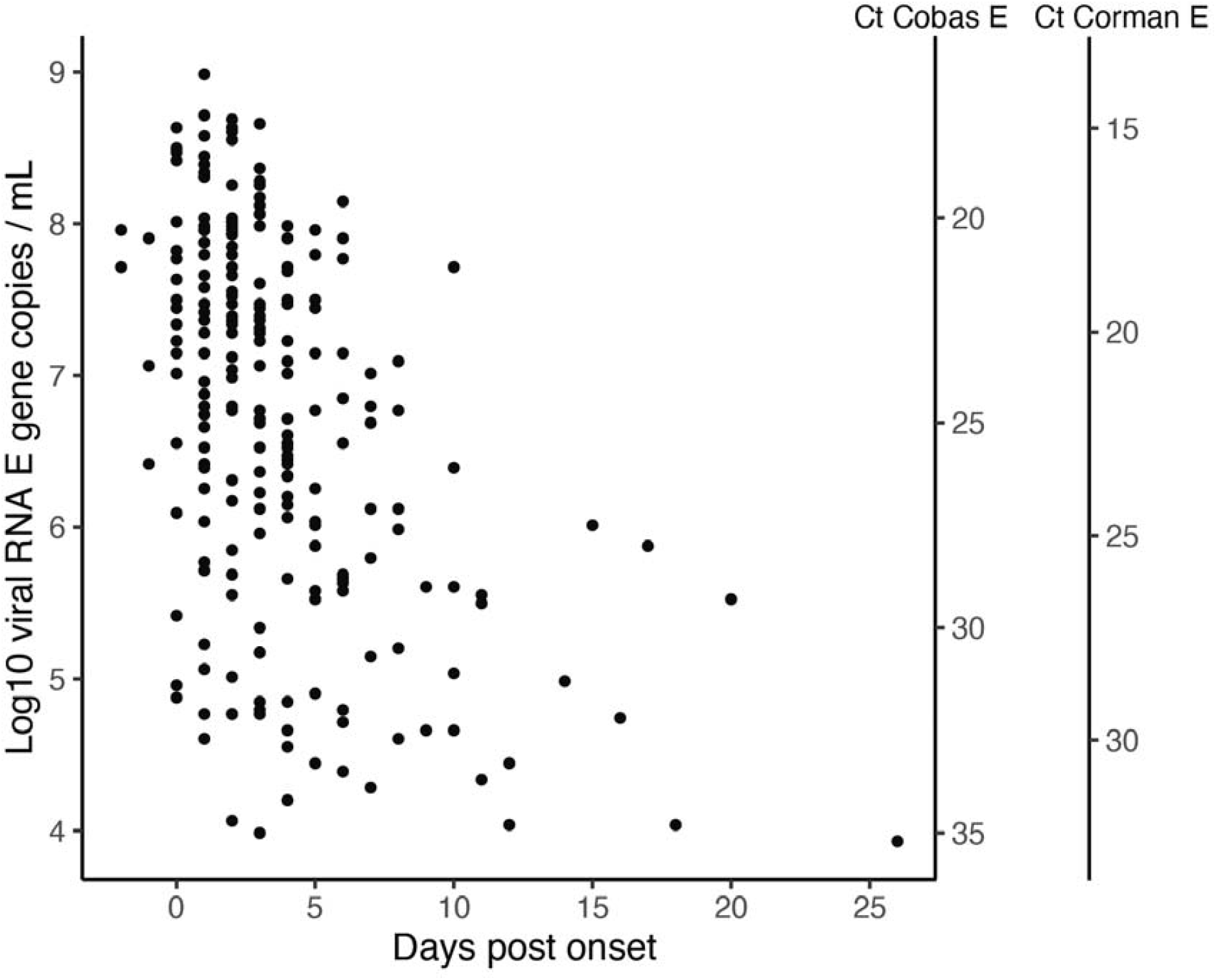
Viral RNA load (left Y axis) by time since onset for n=223 persons testing RT-PCR positive for SARS-CoV-2 at a drive through test station in the city of Rotterdam. Right Y axis shows Ct values for two commonly used RT-PCR platforms.

Table 1 shows the estimated proportion of these infectious cases which would be detected and missed if the different rapid antigen detection tests would be used as the first line of screening in the current test routine. With the most sensitive RDT’s, 97.30% (88.65%-99.77%) of infectious individuals would be expected to be detected. This decreased to 92.73% (range 60.30-99.77) and 75.53 (range 17.55-99.77) for assays B and C, respectively. This preliminary analysis suggests major differences in the RDT’s regarding suitability for tracking of most infectious cases.

**Table 1.** Median, minimum and maximum proportion detected culture positive cases of RT-PCR confirmed cases by rapid antigen tests with different detection limits.

| **Rapid antigen assay** | **Mild, outpatient**<br>**median (min-max)** | **Hospitalised, mild**<br>**median (min – max)** | **Hospitalised, severe**<br>**median (min – max)** |
| --- | --- | --- | --- |
| A | 97.30% (88.65%-99.77%) | 98.68% (95.79%-99.81%) | 99.80% (99.32%-99.97%) |
| B | 92.73% (60.30%-99.77%) | 97.43% (86.40%-99.81%) | 99.54% (97.45%-99.97%) |
| C | 75.53% (17.55%-99.77%) | 91.70% (57.90%-99.81%) | 98.55% (88.53%-99.97%) |

Assay A = Panbio™ COVID-19 Ag rapid test (Abbott), and Standard Q COVID-19 Ag (SD Biosensor);
Assay B = COVID-19 Ag Respi-Strip (Coris BioConcept), and GenBody COVID-19 Ag (GenBody Inc);
Assay C = Biocredit COVID-19 Ag (RapiGEN).

Assuming that the implementation of rapid tests will lead to reporting of the results on the same day, followed by contact by a public health official in all cases, the proportion of cases with optimal start of contact tracing can increase from 33% to 75,53% when using the least sensitive assay to 97.3% for the most sensitive assays.

## Discussion

The use of rapid antigen tests for screening of individuals offers the potential for rapid identification of those individuals at greatest risk of spreading the infection^3^. We tested this line of reasoning based on real life data, as a basis for discussion on choices for assays and testing algorithms. The advantage of faster time to result and therefore initiation of contact tracing is a great added benefit of RDT’s. In our example, routine application of RDT’s will increase the proportion of suspect cases who receive their test results the same day from 33 to 97%. The shortening of testing delays is a critical determinant of success of a contact tracing strategy, and shortening from 3 to 1 days can push expanding outbreaks into suppression with a reproductive number below 1^6,7^. Our analysis also shows, however, that antigen RDT’s differ greatly in their ability to detect infectious cases, therefore requiring careful validation before routine application. In our setting, people were tested relatively soon after onset of disease, when viral loads are at their peak thus ensuring highest sensitivity of the RDT’s. A more challenging application is the use of RDT’s in testing persons without symptoms, as a strategy to reopen society after lockdowns. Here, in the absence of knowledge of time since exposure, negative predictive values are difficult to assess and risk of false negatives is higher than in symptomatic persons that may be using physical distancing as is currently recommended globally. Nonetheless, when used judiciously, RDT’s offer hope to improve containment by more rapid isolation and contact tracing of the most infectious individuals.

## Data Availability

Data is available upon request

## Data Availability

Data is available upon request

## Data Availability

Data is available upon request

## Data Availability

Data is available upon request

## Acknowledgement

This work was funded through EU COVID-19 grant RECOVER (grant agreement ID: 101003589).

