## Supplemental S1 for "From more testing to smart testing: data-guided SARS-CoV-2 testing choices"

**Supplemental file S1**

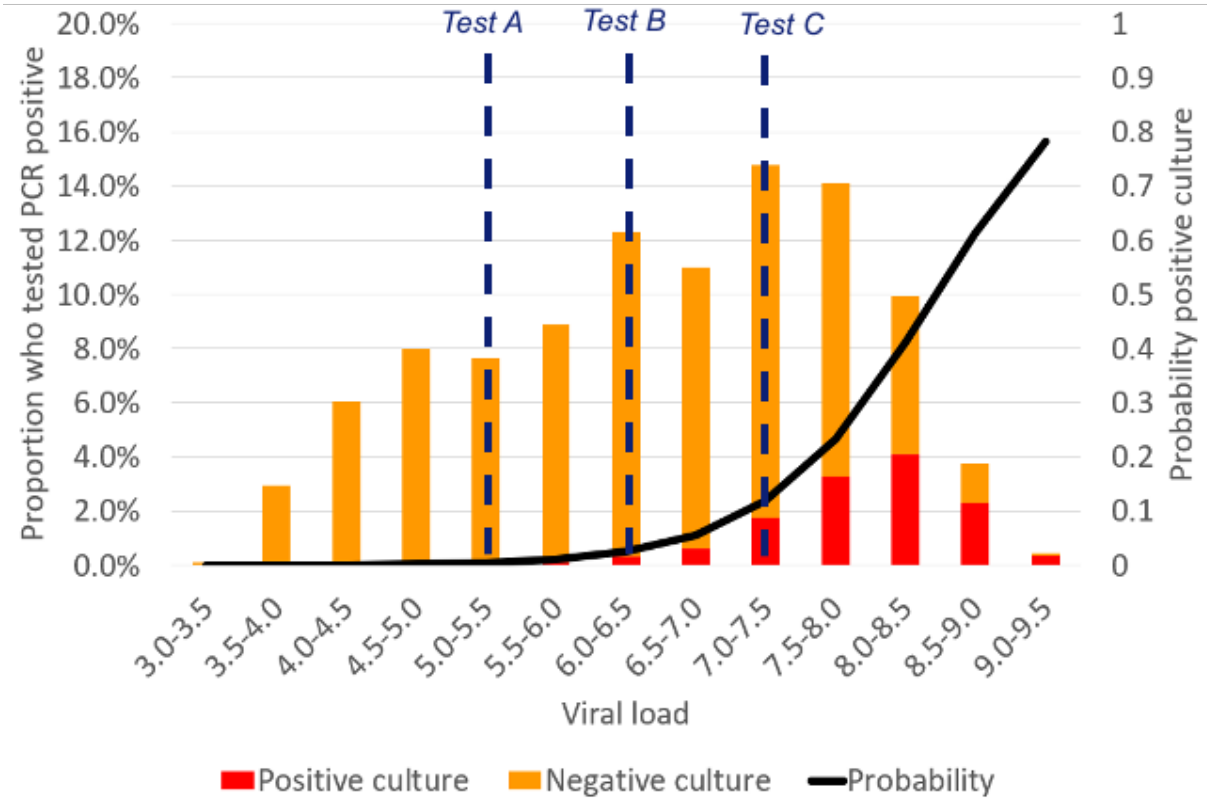


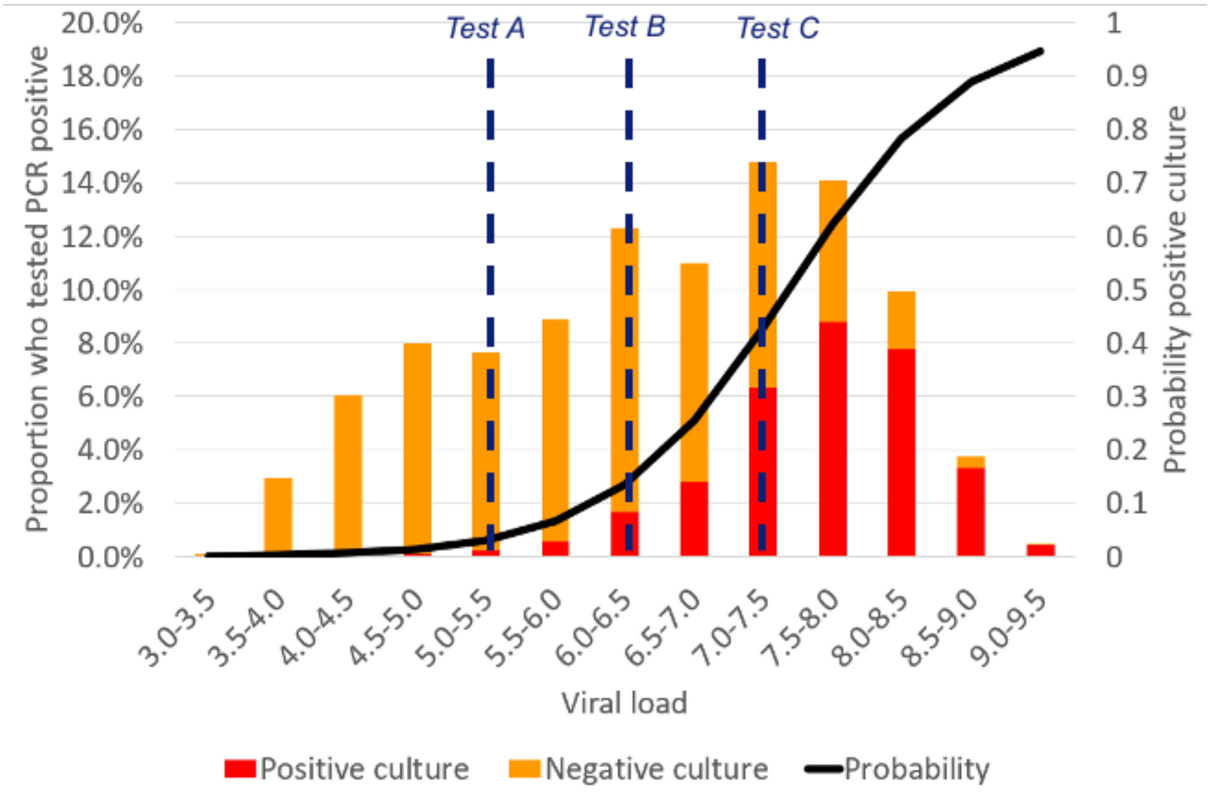

***Supplemental Figure S1****: Distribution of viral RNA loads at time of diagnosis for 1754 patients with SARS-CoV-2 infection presenting to a drive through test station (orange bars), and distribution of patients shedding infectious virus at time of diagnosis (red bars). Black line denotes the projected probability of having infectious virus in the nose/throat swab as calculated using logistic regression based on* **hospitalised severely ill patients** *(Top) (van Kampen et al., 2020) and based on* **hospitalised patients with mild disease** *(Bottom) (Wölfel et al., 2020). Dashed vertical lines indicate the technical detection limits of the different rapid antigen tests.*

*Assay A =* Panbio™ COVID-19 Ag rapid test (Abbott)*, and Standard Q COVID-19 Ag (SD Biosensor);*

*Assay B =* COVID-19 Ag Respi-Strip (Coris BioConcept) *and GenBody COVID-19 Ag (GenBody Inc);*

*Assay C = Biocredit COVID-19 Ag (RapiGEN).*
