## Supplementary material for "From more testing to smart testing: data-guided SARS-CoV-2 testing choices": Online methods

*Patients and metadata*

A database of the municipal health service Rotterdam-Rijnmond with confirmed cases with known onset of disease date detected in June and July 2020 was linked with the laboratory database with viral load data of the Erasmus MC to assess the relationship between viral load and days post disease onset. The distribution of viral load among positive samples collected by the municipal health service Rotterdam-Rijnmond was retrieved from the Erasmus MC laboratory database.

*Diagnostic testing*

Routine RT-PCR testing was performed on combined nasopharyngeal and throat swabs in virus transport medium using the Cobas® SARS-CoV-2 test on the COBAS6800 (Roche diagnostics) or by an in-house developed method (Corman  *et al.*, 2020)) using the MagNApure96 system and LightCycler480 (Roche diagnostics).

*Virus isolation*

For the technical validation of RDTs, SARS-CoV-2/NL/2020 isolate (https://www.european-virus-archive.com/virus/sars-cov-2-strain-nl2020) was propagated by inoculation on Vero E6 cells and harvested after 48 hours. The preparation was not further purified to mimic clinical samples, which contain a combination of released virus particles and cell lysates. Virus stock was titrated in a 10fold serial dilution and inoculated on Vero E6 cells with visual inspection for cytopathic effect (CPE) following 48 - 72 hours (expressed as TCID50). Samples were tested by RT-PCR to determine RNA loads in relation to TCID50 titres of infectious virus^5^. For isolation of infectious SARS-CoV-2 from respiratory tract samples, Vero cells clone 118 was used. Samples were cultured for seven days, and, once cytopathic effect (CPE) was visible, the presence of SARS-CoV-2 was confirmed with immunofluorescent detection of nucleocapsid proteins.

*Genomic copy number calculation*
RT-PCR Ct values were translated into copy numbers by determining the amount of RNA copies based on the E-gene RT-PCR and quantified E-gene in vitro RNA transcripts as described by Corman  *et al.* 2020.

*Antigen rapid tests*

The analytical limit of detection (LoD) of RDT’s was assessed by testing a 10fold serial dilution of cultivated SARS-CoV-2 (both fresh and frozen) in triplicates. samples were 1:1 diluted in the relevant reaction buffer and the recommended amount (i.e. 100 or 130ul), and added to the tests according to manufacture’s instructions. Results were read out after the recommended time. In parallel LoD was also assessed using recombinant purified nucleocapsid from SARS-1 (SARS-CoV N protein, Sino Biological) as it was validated to be very similar to SARS-2^8^. A small set (n=31) of nasopharyngeal swabs in transport media ([HiViral™](http://www.himedialabs.com/TD/MS2760.pdf)) collected from symptomatic people was also tested on the Panbio™ COVID-19 Ag rapid test (Abbott) in parallel with RT-PCR and virus culture.

*Data analyses*

Logistic regression analysis using R version 4.0.0 was used to determine the association between virus positive culture success and 10log transformed viral RNA load. The projected probability for viral infectivity based on viral load was calculated by converting the log odds as calculated in the logistic regression analysis to a probability for viral culture success. We calculated the projected viral isolation success for every viral RNA load found at the time of diagnosis in 1754 confirmed cases diagnosed with SARS-CoV-2 infection presenting at a drive through test station. The number of individuals with a projected viral isolation success was calculated as the sum of the probabilities of viral isolation success times the number of people presenting within a particular viral load range as represented by the bars in Figure 1 and Figure S1.
